# Child stunting from birth to age two years: The Birhan Cohort in Ethiopia

**DOI:** 10.1101/2023.05.20.23290246

**Authors:** Frederick G. B. Goddard, Bezawit Mesfin Hunegnaw, Jonathan Luu, Sebastien Haneuse, Mesfin Zeleke, Yahya Mohammed, Chalachew Bekele, Daniel Tadesse, Meles Solomon, Delayehu Bekele, Grace J. Chan

**Author notes:** **Address correspondence to:** Grace J. Chan, MD MPH PhD, 677 Huntington Ave, Boston, MA 02115. Contributed equally as co-first authors (Frederick G. B. Goddard, Bezawit Mesfin Hunegnaw).

## Abstract

**Objective:** Although there has been a reduction in stunting (low height/length for age), the prevalence of malnutrition in Ethiopia is still high. Child growth patterns and estimates of stunting are needed to determine vulnerabilities and potential for recovery. We collected longitudinal data to determine the prevalence, incidence and reversal of stunting among children aged 0-24 months.

**Methods:** We conducted a cohort study of 4,354 children between December 2018 and November 2020 in the Birhan maternal and child health cohort in North Shewa Zone, Amhara Ethiopia. Children who had their length measured at least once were included in this study.

**Results:** Among the 4,354 children enrolled, 3,674 (84.4%) were included. Our findings indicate that median population-level length is consistently below global standards from birth to age two. Growth velocity was slowest compared to global standards during the neonatal period and after children reached six months of age. The observed prevalence of stunting was highest at age two (57.9%). Incidence of stunting increased over time and reversal was highest between the ages of birth to six months. We found substantial heterogeneity in anthropometric measurements, which we addressed by fitting modeled growth trajectories for each child.

**Conclusion:** Overall, the evidence from this study highlights a chronically malnourished population with much of the burden driven by growth faltering during the neonatal periods as well as after 6 months of age. To end all forms of malnutrition, growth faltering in populations such as that in young children in Amhara, Ethiopia needs to be addressed.

**Article Summary:** Through longitudinal anthropometry data from birth to two years of age, we provide rigorous evidence on the burden, incidence and reversal pattern of childhood stunting.

**What is Known on This Subject:** There is a high prevalence of childhood stunting in Ethiopia. Prior studies show high spatial heterogeneity with highest prevalence in the Amhara region. Ethiopia aims to reduce stunting in children under 5 to meet global targets but remains off track.

**What This Study Adds:** We provide longitudinal data and modeled trajectories to describe patterns of linear growth and onset of growth faltering or recovery. The evidence generated serves as critical input to meet national and global targets of ending malnutrition.

**Contributors Statement Page:** Dr Frederick G. B. Goddard conceptualized and designed the study, curated the data, carried out the initial analyses and interpretation of data, drafted the manuscript and critically reviewed and revised the manuscript for important intellectual content.

Dr Bezawit M. Hunegnaw conceptualized and designed the study, designed the data collection instruments, supervised data collection, contributed to analyses and interpretation of data, drafted the manuscript and critically reviewed and revised the manuscript for important intellectual content.

Dr Sebastien Haneuse and Jonathan Luu contributed to analysis and interpretation of data, and critically reviewed and revised the manuscript for important intellectual content.

Dr Mesfin Zeleke, Yahya Mohammed, Chalachew Bekele and Daniel Tadesse contributed to the design of data collection instruments, supervised data collection and critically reviewed and revised the manuscript for important intellectual content.

Meles Solomon contributed to the interpretation of data and critically reviewed and revised the manuscript for important intellectual content.

Dr Delayehu Bekele oversaw study implementation, contributed to the design of data collection instruments, critically reviewed and revised the manuscript for important intellectual content.

Dr Grace J. Chan obtained funding, conceptualized and designed the study, contributed to analyses and interpretation of data, and critically reviewed and revised the manuscript for important intellectual content.

All authors approved the final manuscript as submitted and agree to be accountable for all aspects of the work.

## INTRODUCTION

One central theme of the United Nations Sustainable Development Goals (SDGs) is child development, including Goals 2 and 4 that aim to end hunger and improve education.^1^ Nutritional status plays a major role in reaching these goals, because it is associated with both physical and cognitive development.^2^ Malnutrition continues to be a major public health problem and is the most important underlying risk factor for illness and death, particularly in low-income countries and among young children.^3^ An estimated 53% of all deaths in children under the age of five are associated with malnutrition.^4^ Target 2.2. under Goal 2 of the SDGs set out to end all forms of malnutrition by 2030, including meeting targets on stunting in children under five by 2025.^5^ Stunting is defined as low length/height-for-age and indicates chronic undernutrition.^6^ Stunting is considered to be an important indicator for both child physical and cognitive development,^7^ and is strongly associated with environmental factors, such as living conditions and nutrition.^8^ In 2019, an estimated 144 million (21%) children under 5 globally were stunted.^9^

To meet the SDGs and end all forms of malnutrition, a wealth of evidence has been generated to understand how and when to intervene. A recent collection of research by the *ki* Child Growth Consortium that combined anthropometric data from over 100,000 children enrolled in 35 longitudinal cohorts across 15 low- and middle-income countries found that the incidence of stunting peaks early between birth and three months.^10,11^ Recovery from stunting was rare, with only 5% of children reversing their stunting status any given month.^11^ The prevalence of child stunting in sub-Saharan Africa remains high, including in Ethiopia.^12^ Although there has been a reduction in stunting over the last two decades, in 2016, an estimated 38% of children under 5 in Ethiopia were stunted.^13^ There is high spatial heterogeneity of stunting within Ethiopia^14^, with the highest prevalence of stunting occurring in the Amhara region.^15^ As part of the World Health Assembly global nutrition targets, Ethiopia aims to reduce stunting in children under 5 to 27% by 2025 with an expected average annual reduction rate of 6%.^16,17^ Current estimates show that the country is off track following an average annual reduction rate of only 2.4%.^18^

To achieve these goals, rigorous studies that not only describe the magnitude of the problem but also the dynamics of growth and changes through childhood are imperative. Accurate depiction of the growth process that includes analysis of growth trajectories and patterns among populations and subgroups is critical to understanding and identifying key time points that predispose children to growth faltering. There are limited longitudinal data on stunting, particularly from underserved populations in low resource settings. While the recent studies by the *ki* Child Growth Consortium included large, pooled datasets of longitudinal anthropometric data, they did not include data from Ethiopia. Infants less than 6 months are often excluded from nutrition surveys and marginalized in nutrition programs,^19^ even though this time period is characterized not only by maximal growth velocity but also by vulnerability to nutrition related events and insults. The Birhan maternal and child health (MCH) cohort in Amhara, Ethiopia, can fill some of these gaps, with longitudinal anthropometry data from birth until the child reaches their second birthday.^20^

## METHODS

### Study design

This study was nested within the Birhan field site, which includes 16 villages in Amhara Region, Ethiopia, covering a mid-year population of 77,766. The site is a platform for community and facility-based research and training that was established in 2018, with a focus on maternal and child health.^21^ Nested in the site is an open pregnancy and birth cohort, the Birhan Maternal and Child Health (MCH) cohort, that enrolls approximately 2,000 pregnant women and their newborns per year with longitudinal follow-up over the first two years of life and household data linked with health facility information.^20^

### Study population

The study population consists of children enrolled in MCH between December 2018 and November 2020, through both a birth and non-birth cohort. The birth cohort included children whose mothers were enrolled during their pregnancy and the child was enrolled at birth. The non-birth cohort includes children that were enrolled after birth at different ages before reaching their second birthday. Children enrolled to the study were followed through age 2.

### Data collection

Custom length board, standardized against the infant/child ShorrBoard (Weigh and Measure, LLC) was used for length measurements during household visits and Seca 417 infantometer was used in the health facilities. Measurement using both models of length measuring devices was recorded to the nearest 0.1cm. Length was measured during child follow-up that was conducted with scheduled visits at birth, on days six, 28 and 42 after birth and at age six, 12 and 24 months, as well as by abstracting facility charts from non-scheduled healthcare-seeking visits (i.e. outpatient visits or hospital admissions).

### Outcomes

The primary outcome of the study was stunting, defined as length-for-age Z-score (LAZ) < -2 SD. We generated length-for-age (LAZ), with length, age and sex data using WHO growth standards.^22^ Z-scores were used to determine the prevalence, incidence and reversal of stunting at each key time point. INTERGROWTH-21 standards were used to generate LAZ scores at birth and during the neonatal period.^23,24^ INTERGROWTH-21 standards were generated using data from low- and middle-income countries and adjust for gestational age at birth, in contrast to WHO standards that assume all babies are born at 40 weeks gestation and hence can overestimate stunting prevalence at birth and during the neonatal period. We excluded anthropometric measurements if 1) their Z-scores fell outside the biologically plausible range defined by the WHO: LAZ< -6 or > 6; or 2) they were abstracted from facility charts after birth, since these measurements were not validated by our data collectors. We compared the median length of children enrolled in the Birhan cohort to World Health Organization (WHO) global growth standards^22^ at key time points. Growth velocity was determined in cm/month between key time points and compared to global WHO standards for the same time periods.^25^

Key time points were defined as birth, four weeks (end of the neonatal period), six weeks, six months, 12 months, and 24 months. Birth length measurements were defined as those that were collected within the first week of life. For measurements at the end of the neonatal period and at six weeks, we included those taken at four weeks and six weeks (+/- one week), respectively, after birth. For the other key time points (six, 12 and 24 months) we included measurements taken within four weeks of each time point.

### Statistical analysis

Empirical length measurements were stratified by sex to describe population median (IQR) length at birth, four weeks (end of the neonatal period), six weeks, six months, 12 months and 24 months of age. Differences in medians between key time points were used to calculate monthly growth velocity. Medians at each key time point and velocities between key time points were then compared to WHO standards. For incidence calculations, the sample population for each time period (defined as two adjoining key time points) was defined as all children that were at risk of stunting (i.e. not already stunted by the beginning of that time period) and have available anthropometric measurements between two adjoining key time points. Similarly, for reversal, the sample population for each time period was defined as all children that had the chance of reversing their stunting (i.e. already stunted by the beginning of that time period) and have available anthropometric measurements at the beginning and end of the time period. To expand on our incidence and reversal analysis, we calculated incidence and reversal both sequentially (i.e. comparing birth to four weeks, then four weeks to six weeks) and using birth as a baseline (i.e. comparing birth to 4 weeks, then birth to 6 weeks).

To consider potential measurement error for length in our findings, we used a mixed effects model to determine potential outliers that should be removed from the analysis. Using LASSO regression to determine significant covariates to include in the models, we considered six different models that used a mix of fixed and random age variables and intercepts, quadratic splines, and piecewise-linear splines. Our final model included a piecewise-linear spline of age for the fixed effect, and an intercept and slope of standardized age for the random effects. We determined this model was the best fit after comparing AIC, BIC, and deviance diagnostic criteria. Using the predicted outcomes and estimated standard deviation from the model, we removed observed points that were not within 1.5 standard deviations of fitted trajectories. After these measurements were removed from the dataset, prevalence, incidence, and reversal estimates were re-calculated. Confidence intervals for all proportion estimates were calculated using the Agresti-Coull method. All analyses were completed in R Version 4.2.3.^26^

## RESULTS

We enrolled 4,354 children under the age of two into the MCH cohort study, among which 3,674 (84.4%) had their length measured at least once and were included in this study (Table 1). Among children included, 82.7% resided in rural areas and 48.7% were female. Among included children with gestational age and birth weight data, 15.3% were born preterm and 9.0% low birthweight. Over the course of the study, length measurements were taken 2.6 times on average for each child, ranging from one to eight measurements for length. Length measurements were clustered around the scheduled follow-up visits (Figure 1).

**Table 1:**
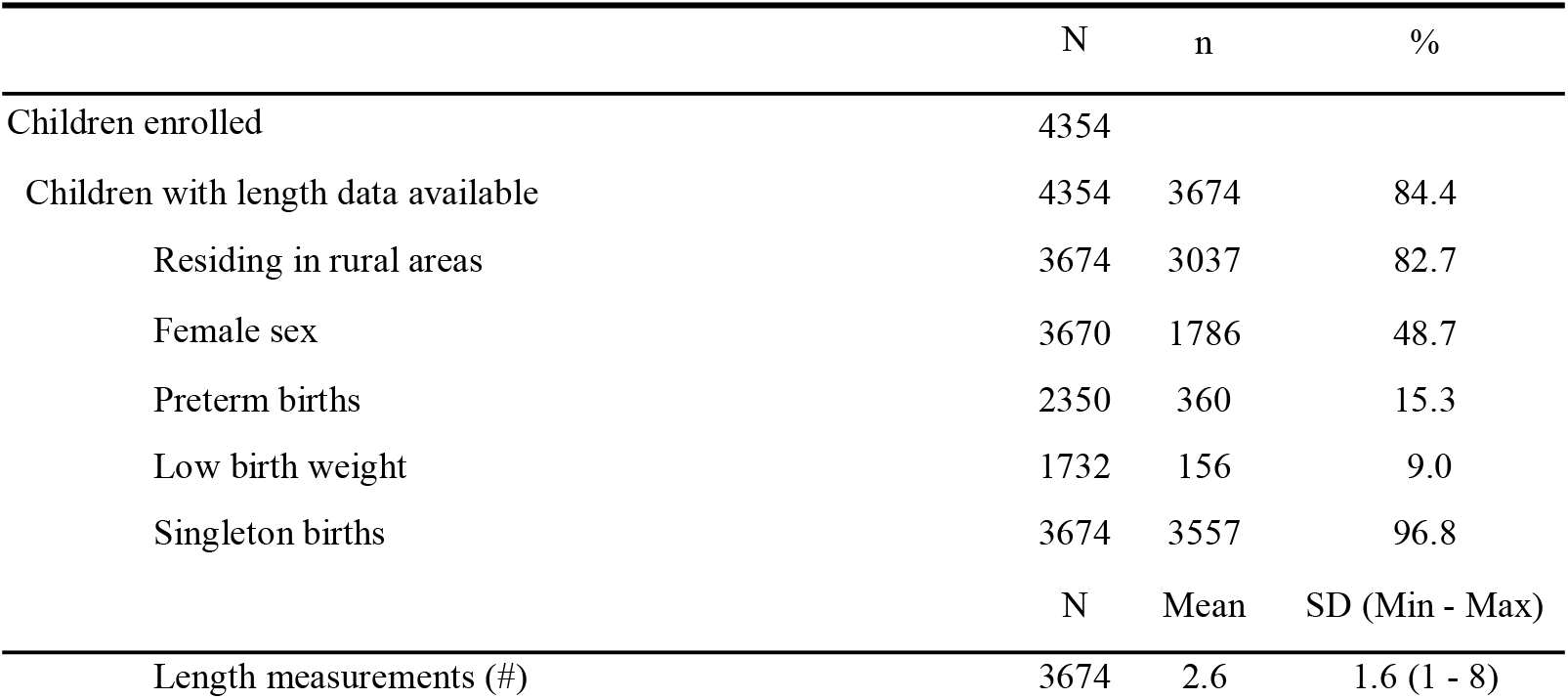
Population summary characteristics

**Figure 1:**
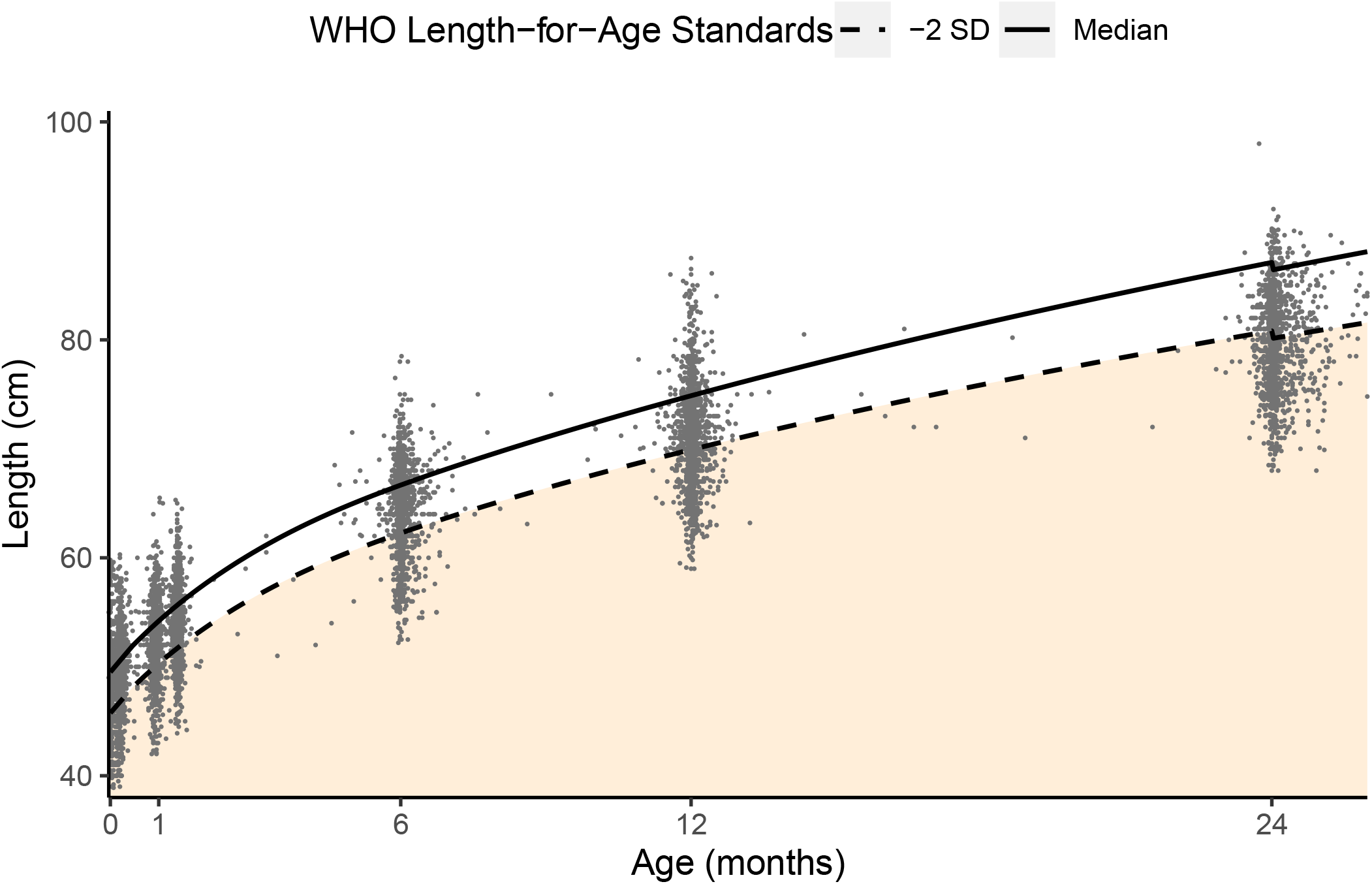
Distribution of length measurements compared to WHO standards (boys and girls combined)

Median length was lower in the study population compared to global WHO standards at all key time points defined for this study, namely birth, the end of the neonatal period (four weeks), at six weeks, and at six, 12 and 24 months (Table 2). The difference was smallest at birth (−0.1cm for girls and -0.9cm for boys), with a slower monthly growth velocity compared to WHO standards thereafter that was most pronounced during the neonatal period (−1.4 and - 1.6cm/month). After the neonatal period, the difference in growth velocity was smaller for girls between weeks four and six and between weeks six and six months (−0.7 and -0.2cm/month).

**Table 2:** Length at key times points for girls and boys compared to WHO global standards

| **Girls** | N | Median (IQR), cm<br>MCH | Median,<br>cm WHO | Difference<br>, cm | Velocity,<br>cm/month<br>Birhan | Velocity,<br>cm/month<br>WHO | Difference<br>,<br>cm/month |
| --- | --- | --- | --- | --- | --- | --- | --- |
| Birth | 882 | 49.0 (47.3-50.2) | 49.1 | -0.1 |  |  |  |
| 4 weeks (end of neonatal period) | 787 | 52.0 (50.0-54.0) | 53.4 | -1.4 | 3.3 | 4.7 | -1.4 |
| 6 weeks | 787 | 53.4 (51.5-55.6) | 55.1 | -1.7 | 3.0 | 3.7 | -0.7 |
| 6 months | 825 | 64.0 (61.5-66.2) | 65.7 | -1.7 | 2.3 | 2.5 | -0.2 |
| 12 months | 863 | 71.0 (68.0-73.5) | 74 | -3 | 1.2 | 1.4 | -0.2 |
| 24 months | 688 | 79.8 (76.4-83.0) | 86.4 | -6.6 | 0.7 | 1.0 | -0.3 |

| **Boys** | N | Median (IQR), cm<br>MCH | Median,<br>cm WHO | Difference<br>, cm | Velocity,<br>cm/month<br>Birhan | Velocity,<br>cm/month<br>WHO | Difference<br>,<br>cm/month |
| --- | --- | --- | --- | --- | --- | --- | --- |
| Birth | 938 | 49.0 (47.5-50.5) | 49.9 | -0.9 |  |  |  |
| 4 weeks (end of neonatal period) | 775 | 52.0 (50.0-54.3) | 54.4 | -2.4 | 3.3 | 4.9 | -1.6 |
| 6 weeks | 778 | 53.8 (52.0-56.0) | 56.2 | -2.4 | 3.9 | 3.9 | 0.0 |
| 6 months | 830 | 65.0 (62.5-67.0) | 67.6 | -2.6 | 2.4 | 2.6 | -0.2 |
| 12 months | 906 | 72.0 (69.0-74.0) | 75.7 | -3.7 | 1.2 | 1.4 | -0.2 |
| 24 months | 733 | 80.0 (76.6-83.0) | 87.8 | -7.8 | 0.7 | 1.0 | -0.3 |

There was no difference in growth velocity for boys in the MCH cohort compared to WHO standards between the end of the neonatal period and age six weeks. Differences in growth velocity were between -0.2 and -0.3cm/month thereafter, resulting in the largest difference between median length in the MCH cohort study compared to WHO standards by age two (−6.6 and -7.6cm).

We investigated the potential effects of measurement error in our data and found variation in longitudinal length measurement for some individuals. To determine which points to exclude in a repeated prevalence, incidence and reversal analysis, we compared the fitted trajectories for each individual with observed data points. Observed lengths that were 1.5 standard deviations away from fitted growth trajectories (n=922) were removed from the modeled prevalence, incidence, and reversal analysis (Figure 2 and Table 3).

**Figure 2:**
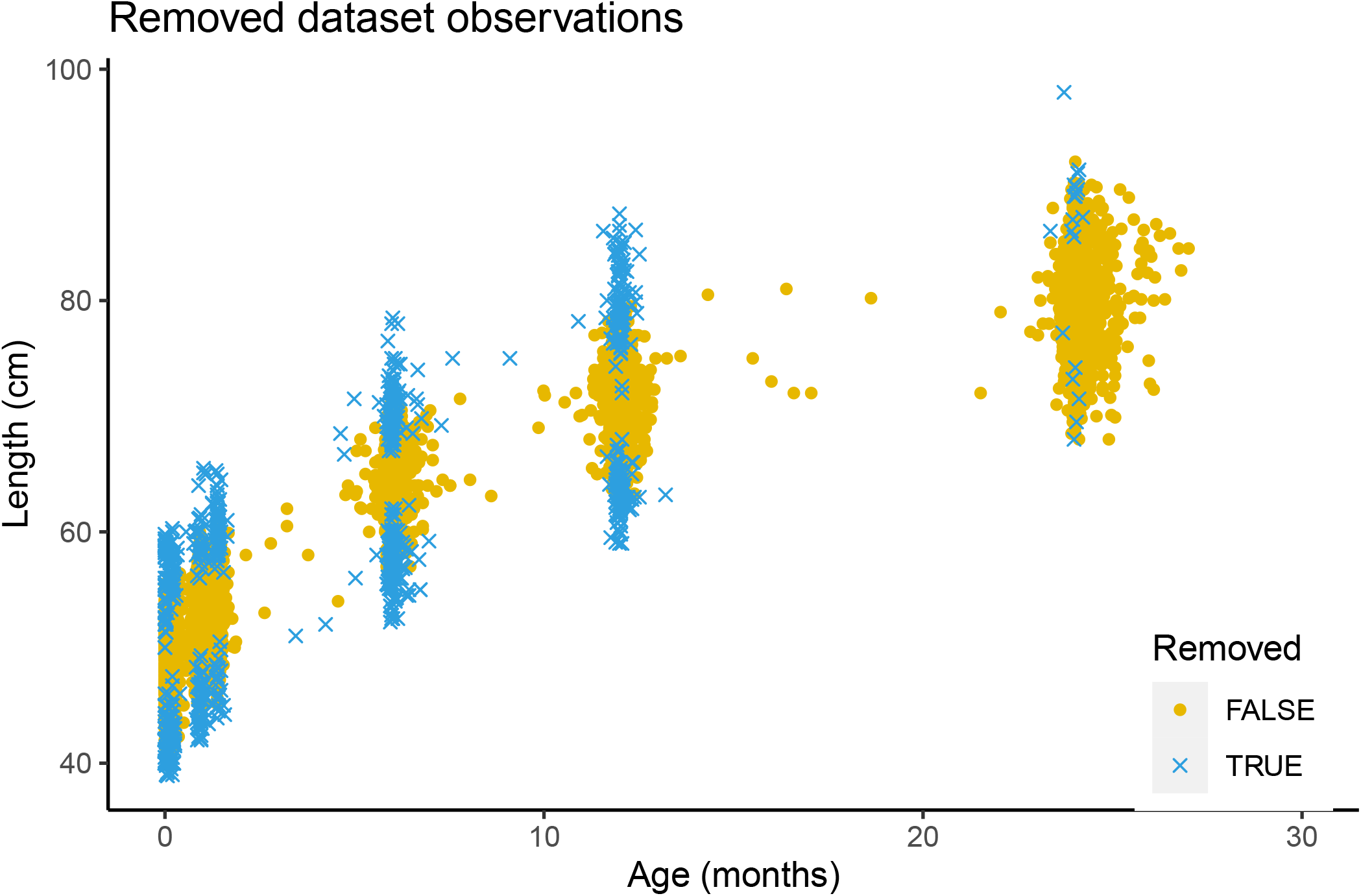
Datapoints kept and removed from dataset for modeled stunting

**Table 3:**
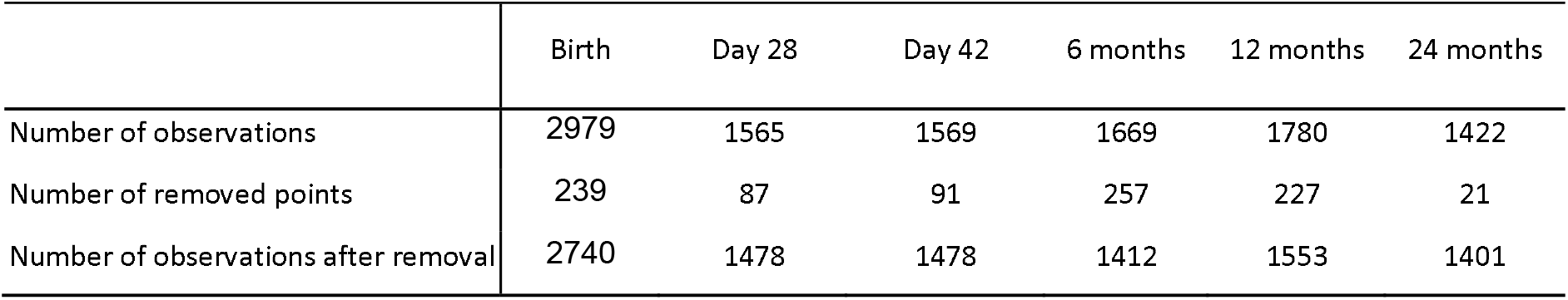
Number of observations

Using observed data, the prevalence of stunting at birth was 16.5% and showed a gradual increase with a sharp rise from the age of 6 to 24 months reaching an observed prevalence of 57.9% among two-year-olds. Observed prevalences from the Birhan data were higher than modeled values at birth and at 6 months, and similar at other key time points(Figure 3).

**Figure 3:**
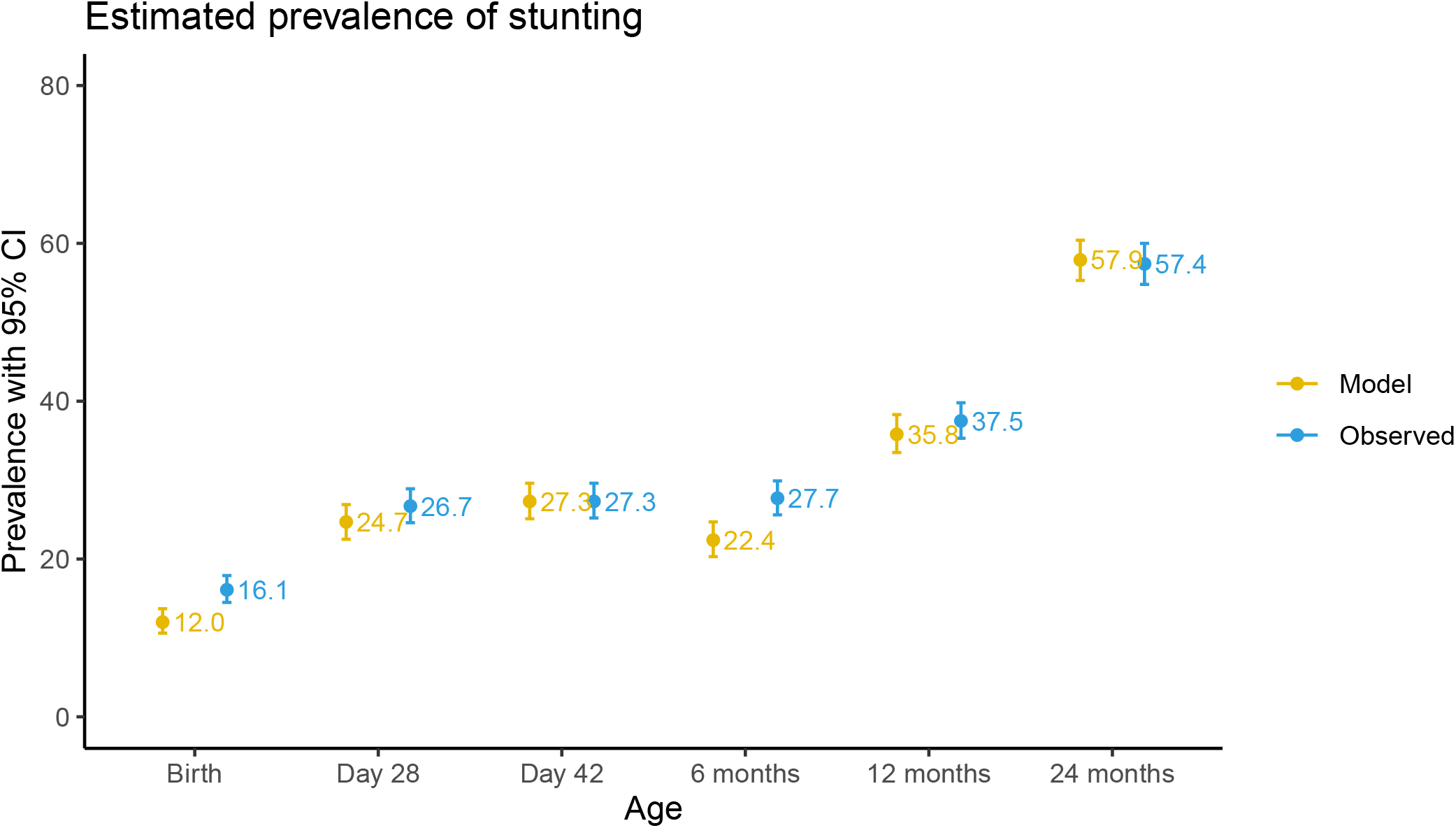
Prevalence of stunting

The incidence, indicating that children were newly stunted at a given key time point compared to the previous time point, followed a similar pattern to the prevalence except for the 6-week timepoint (Figure 4). Among all newborns who were not stunted at birth, 24.4% experienced observed incident stunting within the first month of life. The highest incidence of cases was observed between the ages of 12 to 24 months (51.0%) followed by the 6 to 12 months period (32.5%). Observed incidence estimates were, for the most part, higher than modeled. Further analysis using birth as a common baseline timepoint shows a steady increase in incidence through to the age of two years (Figure S1). Overall, among children who were not stunted at birth, 38.6% had incident stunting at 1 year of age and 56.6% had new onset stunting by 2 years of age.

**Figure 4:**
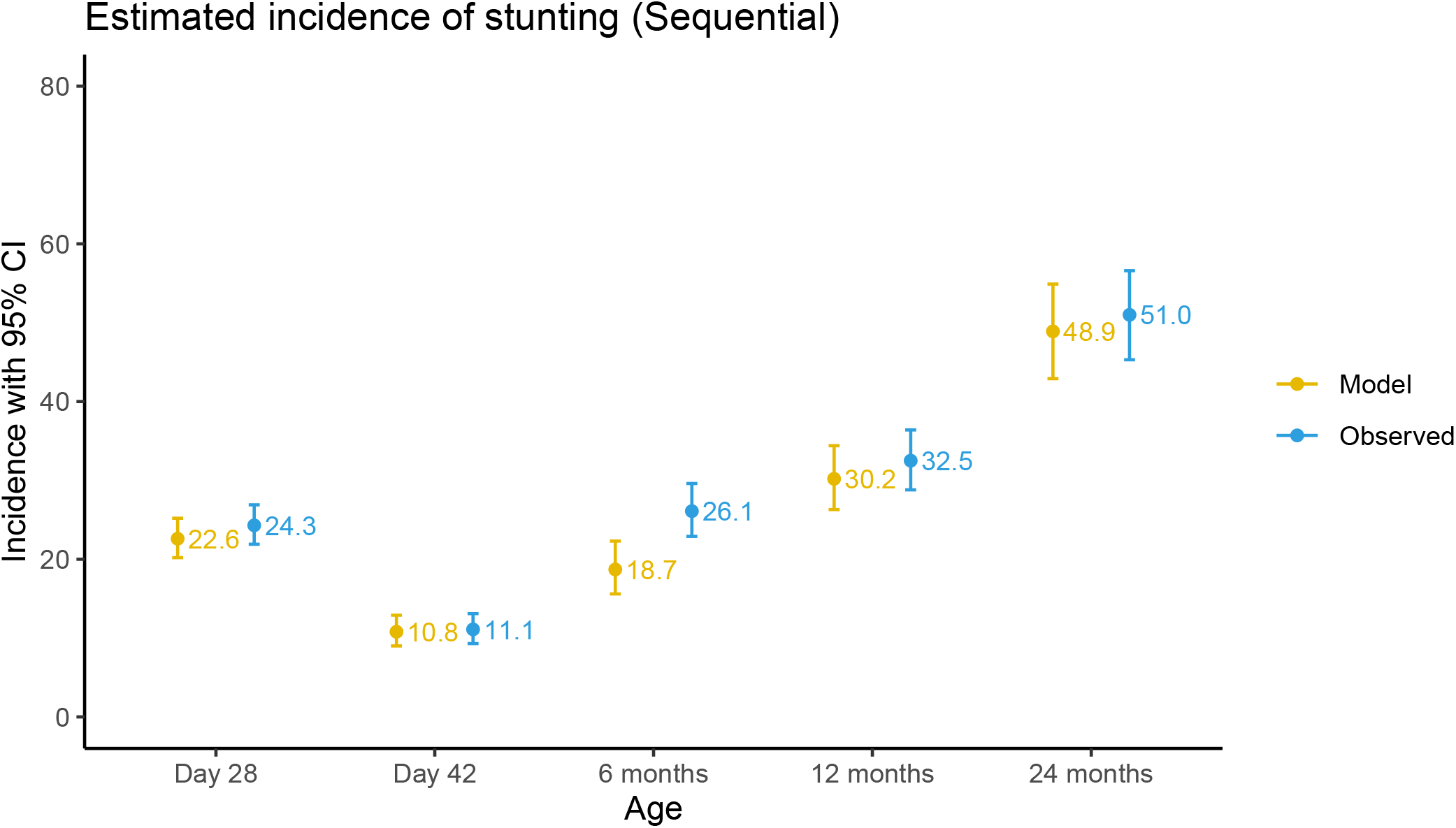
Incidence of stunting (sequential)

The marked difference in incidence between two timepoints compared to difference in prevalence among these same time points indicates the presence of a dynamic incidence and reversal process through time. Rates of reversal (both sequentially when compared to a prior time point and using birth as baseline) were highest by 6 months of age (Figure 5 and Figure S2). Among children who were already stunted at birth and thus had a chance to reverse stunting later in life, 64.1% were able to do so by 6 months of life and 61.5% reversed by the end of the first year of life (Figure S2). By the age of two years, 45.3% of children who were stunted at birth were no longer stunted.

**Figure 5:**
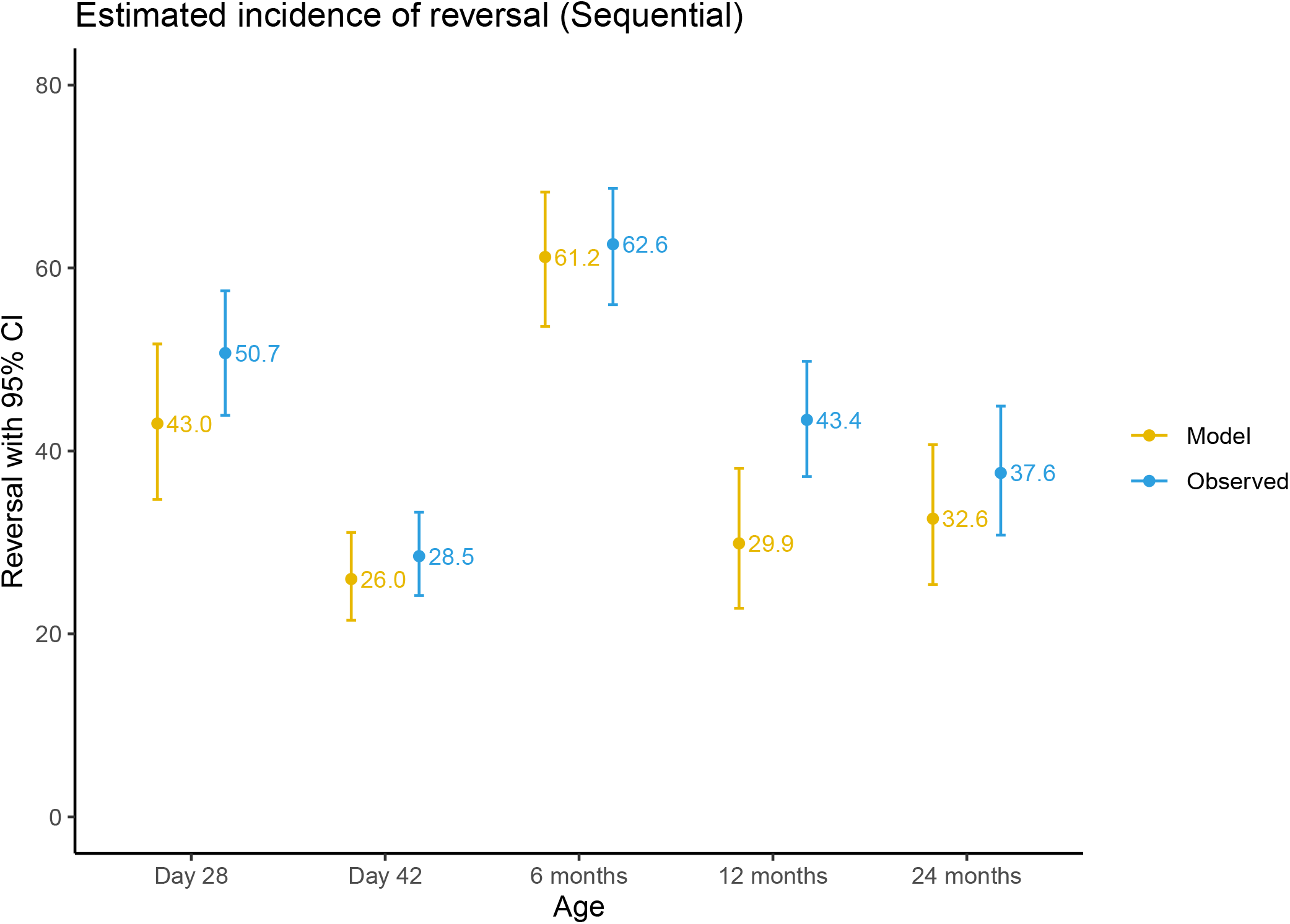
Reversal of stunting (sequential)

## DISCUSSION

Our findings provide evidence that median population-level length among children in our study sample are consistently below global standards from birth to age two. Monthly growth velocity was slowest compared to global standards during the neonatal period, before stabilizing until the child reached six months of age and then slowing again thereafter. Incidence of stunting followed a similar trajectory, suggesting an increasing incidence of newly stunted children over time. Reversal of stunting was a frequent observation with the highest rate of reversal occurring between the ages of birth to 6 months of age.

These findings suggest that among this study population, most loss in potential linear growth is occurring during the neonatal period, and then again, after 6 months of age. An elevated burden in malnutrition after 6 months was also documented in the 2016 Ethiopia Demographic and Health survey (EDHS),^13^ and may be associated with household food insecurity, poor infant and young child feeding practices, lack of food diversity and frequency as well as an increased exposure to enteric pathogens and infectious diseases.^27–29^ The prevalence of stunting at age two in our study was higher than the 2016 EDHS, which found a 47.2% prevalence of stunting among children aged 0-59 months findings in the same region.^15^

Our stunting prevalence findings are consistent with those from the *ki* Child Growth Consortium, that combined longitudinal anthropometric data from multiple cohorts, although did not include data from Ethiopia, where a steady increase was observed in the prevalence of stunting until age two.^11^ However, in contrast to findings from the *ki* Child Growth Consortium, we found that incidence of stunting in this population did not peak during the first three months of life, but rather continued to increase throughout the first two years of life. Stunting reversal was much higher in our study population, particularly early in life from birth to 6 and 12 months of age.

This may be reflective of the impact of various initiatives in place throughout the country to effect reduction of stunting, which include nutritional interventions, improved health care access, sanitation and education.^30^ Additionally, the infancy period is also marked by the greatest potential increment in linear growth, thereby presenting higher likelihood of favorable interventions to effect reversal changes. However, the high prevalence and incidence of stunting suggests that even when stunting status is reversed, a substantial number of children relapse.

This study also found evidence of within-child heterogeneity in longitudinal length measurements that may be due to measurement error. Challenges associated with measuring length among newborns, infants and toddlers have been well documented.^31,32^ In this study, we excluded measurements that were not biologically plausible as defined by WHO standards. We also developed modeled trajectories with previously validated methodologies using linear spline multilevel models that have knots at key time points,^33–35^ to validate the observed prevalence, incidence and reversal findings. Excluding outliers did not substantially alter prevalence, incidence or reversal findings. Observed values were slightly higher than modeled estimates especially in the earlier ages implying that our observations may have overestimated the outcome, possibly related to difficulty in measuring length in the younger ages.

The findings of this study should be interpreted with its limitations in mind. We saw heterogeneity of length measurements for individual children, both at the same time point and across two time points. We were also limited in the number of measurements we had per child and each child contributed a different number of observations to this analysis due to late enrollments in the non-birth cohort, intermittently missed scheduled visits, differences in the prevalence of unscheduled healthcare-seeking visits, or missing data from the pausing data collection due to COVID-19. We also recognize that incidence and reversal events or relapses may occur within a single longer study time frame, which may have been identified with shorter time windows especially in the post infancy period. However, this study provides several important contributions to the literature. It outlines longitudinal evidence on the burden, onset and reversal of growth faltering in Amhara, Ethiopia, the region identified by the 2016 EDHS as having the highest burden of malnutrition in the country.^13^ This study also provides evidence of linear growth at birth and during the neonatal period, where available data are more limited.

Finally, there is growing recognition that growth faltering is a dynamic process with the burdens of undernutrition not limited to those that pass the threshold for stunting at a defined point in time.^6^ Instead, it is worth noting, that the evidence from this study suggests that this population is chronically malnourished compared to global standards. To meet the SDGs and end all forms of malnutrition, then growth faltering in populations such as that in children under the age of two in North Shewa Zone, in Amhara, Ethiopia needs to be addressed.

## Supporting information

Incidence of stunting (birth as common baseline)

Reversal of stunting (birth as common baseline)

## Data Availability

Data are available upon reasonable request.

## Abbreviations

SDGs: Sustainable Development Goals
MCH: Maternal Child Health
LAZ: Length-for-Age Z-score
WHO: World Health Organization
AIC: Akaike Information Criteria
BIC: Bayesian Information Criteria
EDHS: Ethiopia Demographic and Health Survey

## ACKNOWLEDGEMENTS

We would like to thank all mothers and children who participated in the study and the community of the Birhan field site. We are grateful for our partnership with the Ministry of Health, St. Paul’s Hospital Millennium Medical College, Amhara Regional Health Bureau, North Shewa Zone Office, Angolela Tera and Kewet/Shewa Robit Woreda Health Offices and catchment health facilities. We thank the data collectors, supervisors, coordinators and the HaSET team for their significant contributions.

## Supplementary material

Figure S1: Incidence of stunting (birth as common baseline)

Figure S2: Reversal of stunting (birth as common baseline)

