## Supplementary figures and images for "Child stunting from birth to age two years: The Birhan Cohort in Ethiopia"

### Incidence of stunting (birth as common baseline)

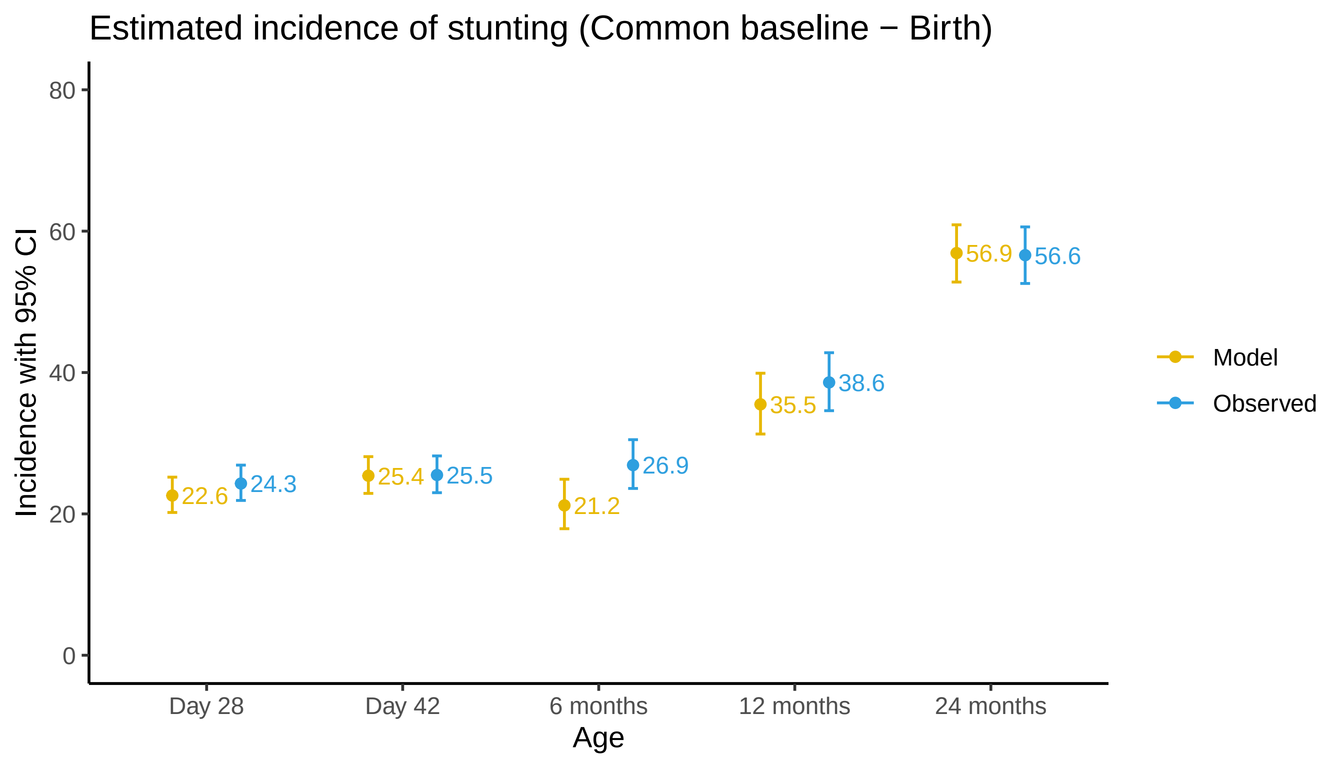

### Reversal of stunting (birth as common baseline)

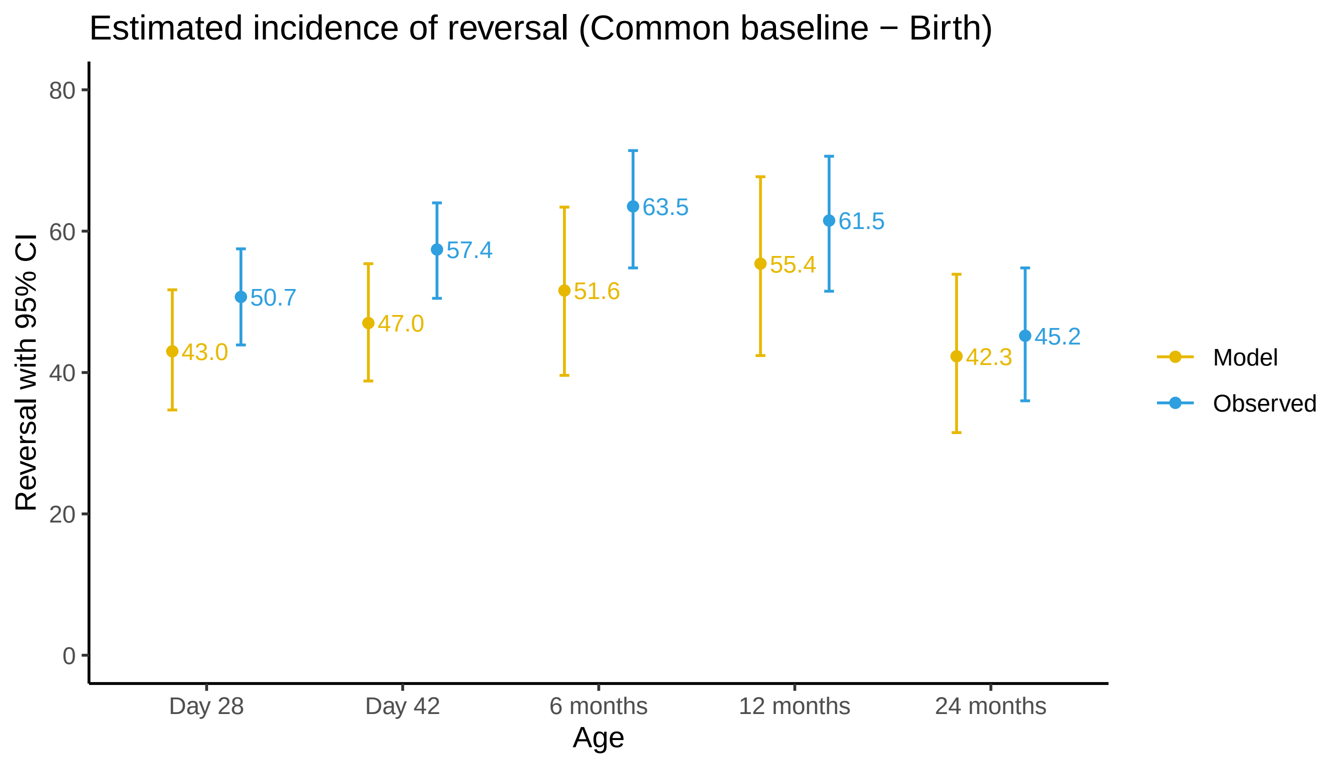
